# Proteome-wide Mendelian Randomization Analysis Identified Potential Drug Targets for Myocardial Infarction

**DOI:** 10.1101/2023.12.05.23299535

**Authors:** Xufeng Ye, Xinpei Wang, Jinzhu Jia

## Abstract

**Background:** Myocardial infarction (MI) is a leading cause of global mortality. Finding effective drugs to treat MI is an urgent concern for clinicians. Proteome-wide Mendelian randomization (MR) analysis provides a new way to investigate invaluable therapeutic drug targets more efficiently.

**Methods:** Using a proteome-wide MR approach, we assessed the genetic predictive causality between thousands of plasma proteins and MI risk. First, by adopting several principles to judge genetic variants associated with plasma proteins and MI risk, we selected a series of suitable variants utilized as instrumental variables (IVs) for the latter Mendelian Randomization (MR) analysis. Second, we performed a proteome-wide MR analysis to select candidate proteins. Third, sensitivity tests including heterogeneity test, reverse causality test, and colocalization analyses were conducted to ensure the robustness of our selected protein. Last, we assessed the drugability of the identified potential drug targets for MI using databases including DrugBank, PharmGKB, and TTD.

**Results:** Of the identified IVs, 3,156 associated with 1,487 plasma proteins were validated. 15 proteins exhibited significant genetically predicted causal associations(P − value < 3.362∗10^−5^) with MI risk, including Plasmin, MSP, Apo B, TAGLN2, LRP4, C1s, Angiostatin, Apo C-III, PCSK9, ANGL4, FN1.4,Apo B, IL-6 sRa, SWAP70, FN, FN1.3. Sensitivity analyses pinpointed Plasmin and Angiostatin for heterogeneity and proteins MSP, Apo B, and Angiostatin for reverse causality effects. Colocalization analysis found several proteins sharing genetic variants with MI, notably Apo B, TAGLN2, LRP4, C1s, Apo C-III, PCSK9 and ANGL4. When the threshold was lowered to 0.7, additional variants SWAP70 could be contained. 7 potential drug targets for MI were identified: SWP70, TAGLN2, LRP4, C1s, Apo C-III, PCSK9, and ANGL4. Drugability assessment categorized these proteins into varying therapeutic potential categories, from successfully drugged targets to those only reported in the literature.

**Conclusion:** Our comprehensive study elucidated 7 promising drug targets offering profound insights into its molecular dynamics and presenting potential pathways for therapeutic interventions against MI.

**Clinical Perspective:** *1) What Is New?:* ★ The analysis of thousands of proteins has identified 7 proteins that have a potential causal role in myocardial infarction risk. ★ Four of these ten proteins have drugs approved or in development that target them, and three 5 have not been previously reported to be associated with atrial fibrillation risk.

*2) What Are the Clinical Implications?:* ★ The results of the present study demonstrate new potential drug/therapeutic targets for myocardial infarction.

## Introduction

Myocardial Infarction (MI) ranks among the primary cardiovascular diseases world-wide, which mainly originates from insufficient or interrupted blood supply via the coronary arteries, leading to cardiac muscle damage or necrosis [1]. Despite advancements in risk factor management and pharmaceutical interventions aiming to alleviate the burden of MI, its incidence and prevalence have steadily surged in the past two decades and will be rising in the forthcoming decades [2]. Contemporary therapeutic strategies, encompassing both extant and drugs under development, often come under scrutiny due to concerns regarding their safety and efficiently [3]. Consequently, the pursuit for more efficient and safe drug targets is at the forefront of clinicians and researchers’ priorities.

### Proteomic Considerations in MI Pathophysiology

Proteins play a vital role in various biological processes at the cellular level, establishing themselves as crucial components for potential drug targets [4–6]. Numerous studies have underscored a significant correlation between levels of various plasma proteins and the risk of MI [7–9]. However, the inherent constraints of observational research, such as residual confounding and potential reverse causality, render these significant observations non-representative of clear causal relationships [10, 11].

### Advancements in Cardiovascular Risk Factor Targeting

Cardiovascular risk factor identification and targeted causal inference have made remarkable strides in the prevention and treatment of coronary artery diseases, including MI. Propelled by modern medical advancements, epidemiological studies have investigated hundreds of biomarkers (circulating molecules, physiological traits, and other pathologies) and targets linked with coronary artery disease. [12, 13] The potency and replicability of associations, coupled with the inherent mechanisms delineated by foundational medicine, have led researchers to causally link certain novel biomarkers and targets with MI [14, 15]. Subsequent interventions aiming to prevent these identified “risk factors” have been initiated. However, differences often emerge between these findings and those observed in epidemiological settings [16]. The ambiguity surrounding the causal roles of numerous biomarkers during disease progression together with the steep costs of drug development programs reveals the necessity to distinguish between “causality” and mere “association” [17].

### Mendelian Randomization (MR) as a Research Tool

MR offers a robust statistical apparatus, furnishing a unique approach to investigating causal relationships between exposures and outcomes [18, 19]. By using genetic variations as instrumental variables to estimate the causal effects of plasma proteins on MI, we can obtain more precise causal estimates than conventional observational studies [14–17]. Notably, MR studies on plasma proteins and MI predominantly feature a few proteins grounded in extant knowledge, sidelining a variety of other proteins potentially correlated with MI risk.

### Research Aims and Methodology

This study endeavors to leverage the MR framework to holistically analyze potential causal relationships between thousands of plasma proteins and MI risk, streamlining the identification of novel MI drug targets. Our methodology incorporates the selection of independent genetic loci, significantly correlated with protein levels and proximate to protein-encoding gene locations, as instrumental variables, substantially mitigating the possibility for violating instrumental variable assumptions. We further fortify our results through a series of sensitivity analyses. We anticipated that this investigation will shed comprehensive insights, serving as a cornerstone for MI drug target research and development.

## METHODS

### Data Source And Disclosure Statement

The database for genetic variations associated with plasma proteins has been sourced from deCODE genetics/Amgen, Inc., Reykjavik, Iceland. The database for genetic variations associated with myocardial infarction (MI) has been obtained from two major repositories: the UK BioBank and the CARDIoGRAM+C4D Consortium. Both of these databases are publicly accessible, in accordance with data sharing policies.

### Discovery analysis

#### Mendelian randomization

Figure 1 illustrates the working principle of Mendelian randomization. Firstly, we define certain mathematical symbols as depicted in the figure. We denote the independent instrumental variables (IVs) as gi. Here, X represents exposure, Y stands for the outcome, and U signifies the confounders. The aggregate effects of gi on X and Y are represented as β_Xi_ andβ_Yi_, respectively.

**Fig. 1.**
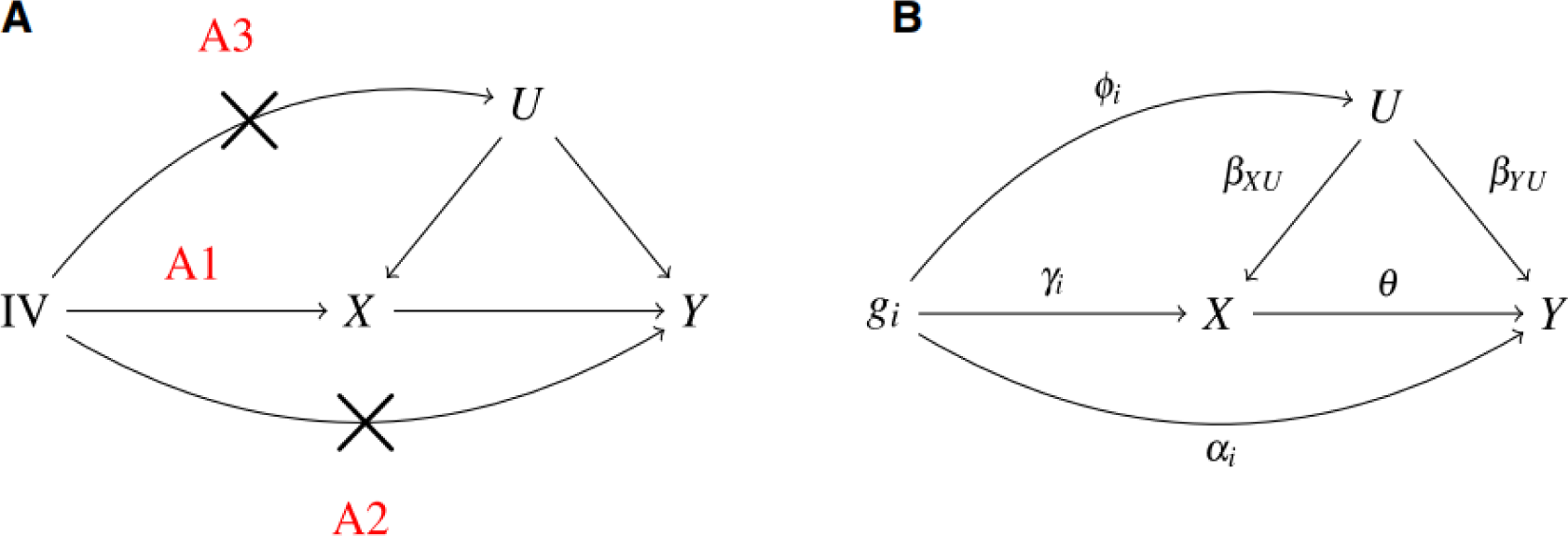
The working principle of Mendelian randomization

**Fig. 2.**
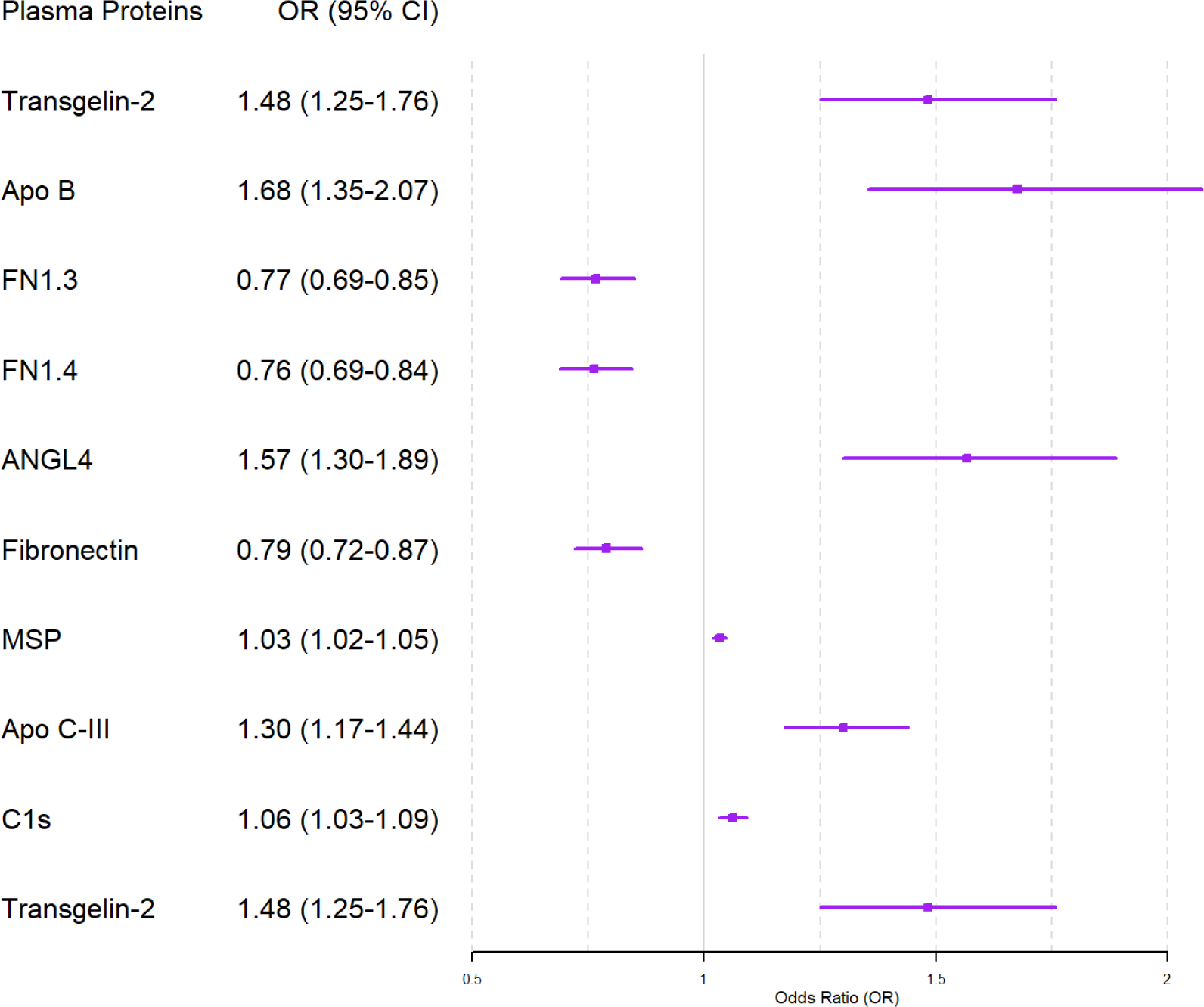
Results of selected proteins with single snp

**Fig. 3.**
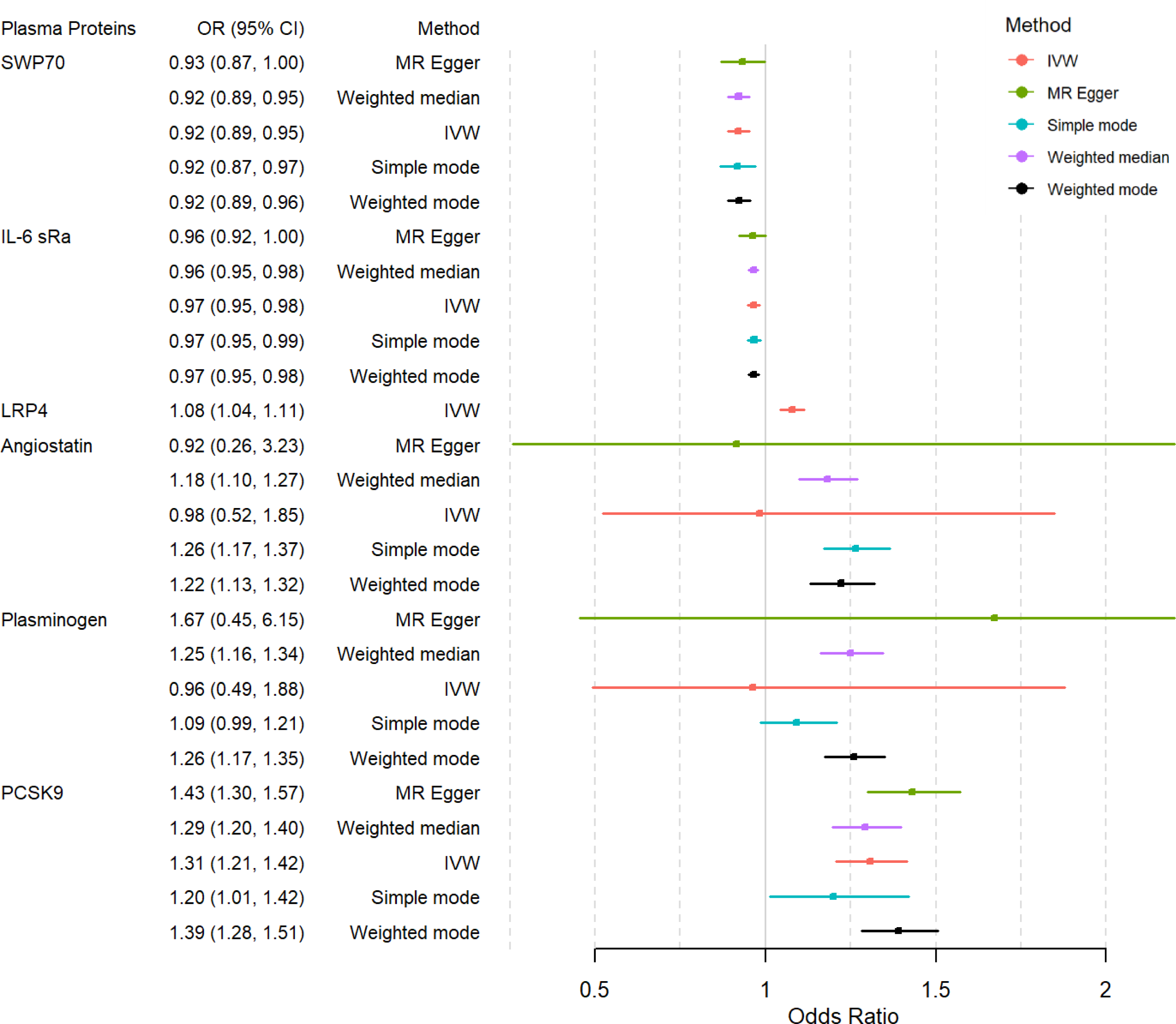
Results of selected proteins with several snps

To avoid the potential for biased inference from horizontal pleiotropy (where IVs affect Y not just through X), Mendelian randomization requires three core instrumental variable (IV) assumptions [20]:

(A1) Relevance assumption: The genetic variants used as instruments are associated with the exposure. Mathematically, the IV is correlated with exposure X; that is, *γ*_*i*_ ≠ 0.

(A2) Independence assumption: The genetic variation and the outcome of interest share no common causes (i.e., no confounders). Mathematically speaking, the IV is uncorrelated with the outcome Y given the exposure X; that is, *α*_*i*_ = 0.

(A3) Exclusion restriction assumption: There are no independent pathways between the genetic variation and the outcome apart from through the exposure. Mathematically, the IV is uncorrelated with unmeasured confounders U; namely, *φ*_*i*_ = 0.

Given the above assumptions, we have equations (1) and (2): Under the circumstance of assumption (A2) and (A3), *α*_*i*_ = 0 φ_*i*_ = 0. Then, we collect causal effect *θ*= *β*_Y*i*_ / *β*_X*i*_, which is called wald ratio.

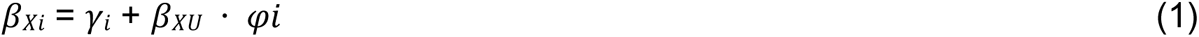

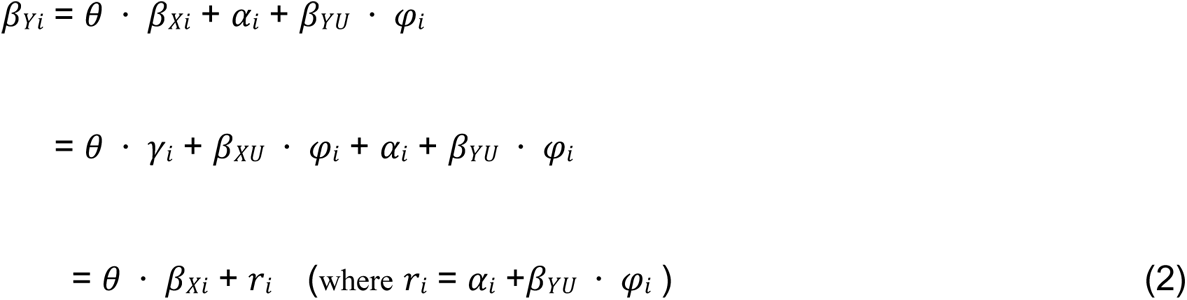

### IV Selection and Validation

Genetic variations utilized as IVs need to adhere to the three assumptions previously stated. The first of these assumptions postulates a correlation between genetic variations and the exposure, (various types of plasma proteins). To substantiate this assumption, we drew upon the findings of a genome-wide association study (GWAS) conducted by Ferkingstad, E. et al[21]. This study delved into the associations of 27.2 million genetic variants with 4,907 aptamers, quantifying 4,719 plasma proteins in a sample of 35,559 Icelanders (courtesy of the Icelandic Cancer Project and deCODE genetics, Reykjav′ık, Iceland). Subsequent to this, we applied rigorous filters to this pool of pQTLs, eliminating: 1. Non-biallelic single nucleotide polymorphism (SNPs); 2. SNPs with minor allele frequency (MAF) < 0.01; 3. Variants located either on the sex chromosomes or within the human major histocompatibility complex (MHC) region.

To satisfy Assumptions 2 and 3, we employed more stringent criteria in selecting pQTLs: 1. Only pQTLs located within 200kb upstream of the transcription start site or 200kb downstream of the termination site of their corresponding protein-coding genes were included and labeled as cis-pQTLs; 2. For these cis-pQTLs, we counted the number of proteins associated with each pQTL, as well as those linked to this pQTL in linkage disequilibrium (LD R2 > 0.5). We then excluded pQTLs that had associations with more than five proteins. It’s worth noting that our criteria for selecting cis-pQTLs are stricter than those in many standard studies. Our primary aim is to ensure the instrumental variables (IVs or pQTLs) we choose influence the outcome (MI) solely through their corresponding proteins, thus minimizing the impact of horizontal pleiotropy on our MR estimates.

Finally, to mitigate the impact of correlation between IVs on the MR estimates, we clumped the above-selected IVs (pQTLs). In alignment with Albert’s research, we controlled LD R2 between IVs below 0.4[22]. This threshold is backed by evidence suggesting that the issue of unstable coefficient estimation due to collinearity begins to manifest when the LD R2 between independent variables exceeds 0.36[23]. Subsequently, we assessed the strength of each IV for every protein. Proteins with insufficient IV strength (with an F − statistic < 10) were excluded, ensuring that the IVs we employed effectively represented genetic proxies of protein levels. Through these rigorous quality control measures, we ensured our chosen IVs for MR analysis faithfully adhered to the foundational three assumptions of Mendelian Randomization.

### Statistical analysis

For our primary MR analysis, we utilized the IVs shortlisted from the aforementioned procedures. The data for our outcome variable, MI, was extracted from a GWAS encompassing approximately 831,000 individuals of European ancestry, an amalgamation of several contributory cohorts. Within this, the UK Biobank provided data on 17,505 MI cases and 454,212 controls. It pinpointed 1,966 single-nucleotide polymorphisms (SNPs) spread across 31 loci, all exhibiting associations with MI and surpassing the genome-wide significance threshold (P=5∗10^−8^). In contrast, the CARDIoGRAM+C4D Consortium contributed information on 61,000 MI cases, 578,000 controls, and a sweeping range of 8,126,035 SNPs. Of these, 4,419 variants, dispersed over 80 loci, were deemed significant.

In scenarios where only one IV corresponded to a protein, we employed the wald ratio method to estimate the causal effect. However, for proteins linked to two or more IVs, we use MR-Egger [24], the inverse variance weighted method (IVW) [25], weighted mode method [26], simple mode method [24] and weighted median method [27] for our MR estimation. To ensure precision, we use Bonferroni correction to adjust the MR results. Associations reflecting a P −value < 0.05/ (the number of proteins assessed in MR) were classified as significant. We use R package “TwoSampleMR” (https://github.com/MRCIEU/TwoSampleMR) to execute MR analysis.

### Sensitivity analysis

#### Horizontal Pleiotropy Test

Horizontal pleiotropy arises when a genetic variation affects the outcome via path-ways other than the primary exposure, which can introduce biases into previously calculated MR estimates. To address this, we employ Egger regression [28] to assess the presence of horizontal pleiotropy. Theoretically, in the absence of horizontal pleiotropy, IVs impact the outcome solely through the exposure. Within this regression model, the slope quantifies the effect of exposure on the outcome, while the intercept serves as an indicator for pleiotropy. A significant deviation of the intercept from zero signals the existence of horizontal pleiotropy.

#### Heterogeneity Test

A fundamental assumption of Mendelian randomization (MR) is that genetic variations correlate with the exposure but are independent of any confounding factors that could bias the causal association between the exposure and the outcome. However, the genetic variations utilized as instrumental variables (IVs) might sometimes violate this assumption. This can occur if they are related to other factors influencing the outcome or different genetic variations impact the exposure through diverse pathways. In MR analysis, the heterogeneity test serves as an essential tool to assess if the genetic variations used as IVs align with the MR assumptions.

A widely employed method to conduct this heterogeneity test is Cochran’s Q test[29]. The specific steps are:

1. For each genetic variation, define 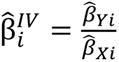 with an approximate standard error given by 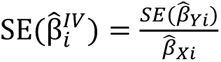, where 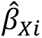 and 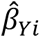 represent the effects of the IV on the exposure and outcome, respectively.
2. Compute the statistic 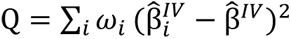, where 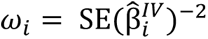 is the inverse variance weight and 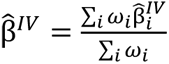 is the inverse variance-weighted estimate based on all genetic variations (fixed effects).
3. Under the null hypothesis, the statistic Q follows a chi-squared distribution *X*^2^(I − 1). If this is significant, it indicates heterogeneity among the genetic variations, suggesting potential violations of MR assumptions.

#### Reverse causation

Although the motivation of Mendelian randomization is to avoid bias from unmeasured confounding and reverse causation, the method may invalidate causal conclusions in several circumstances [30]: Genetic association with the risk factor is not primary, Feedback mechanism and Cross-generational effects. To enhance the robustness of our Mendelian randomization, we implemented MR-Steiger method [31] to undertake a directionality test. Additionally, we conduct a reverse Mendelian Randomization to verify whether there exists causation from MI to plasma proteins, which utilize MI as an exposure and plasma proteins as outcome.

#### Colocalization

Proximal variants often inherit together and therefore are usually correlated, which is termed as linkage disequilibrium law in genetics. Two traits might be causally influenced by distinct variants that happen to be correlated, providing a pathway between genetic variation and an outcome not mediated by the exposure, which could violate the exclusion restriction assumption of Mendelian randomization. We employ colocalization analysis to determine whether a specific genetic locus’s variant is associated with both a plasma protein (a particular exposure) and MI (a health outcome). The central premise of colocalization analysis is to determine whether a particular variation at a genetic locus is associated with both an exposure and a health outcome. We use R package “coloc” (https://github.com/chr1swallace/coloc) to execute colocalization analysis.

### Potential Drug target

To investigate the current state of drug development focused on these identified proteins, we consulted three specialized databases: the DrugBank Database, the PharmGKB Database, and the Therapeutic Target Database (TTD), each with its own unique features. The DrugBank Database [32] offers comprehensive data on FDA-approved as well as experimental drugs, including details on drug targets, mechanisms of action, and drug interactions. PharmGKB[33], provides international drug labeling information over various countries, including the United States, Canada, Europe, Switzerland, and Japan. The TTD Database [34] offers valuable literature-based information on known therapeutic protein and nucleic acid targets, targeted disease conditions, pathways information and the respective drugs or ligands for each target.

### Ethics statement

Patients or the public were not involved in the design, conduct, reporting or dissemination plans of our research.

## Results

### IV selection and validation

Ferkingstad, E. et al’s research delved into the associations of 27.2 million genetic variants with 4,907 aptamers, quantifying 4,719 plasma proteins in a sample of 35,559 Icelanders (courtesy of the Icelandic Cancer Project and deCODE genetics, Reykjavik, Iceland). From this extensive dataset, we earmarked 4,631 proteins as potential exposures and identified 28,191 pQTLs associated with these proteins (P−value < 1.8∗10^−9^, (0.05/27.2 million)) as candidate IVs for our Mendelian Randomization (MR) analysis. After excluding IVs to satisfy the assumption of MR analysis, we select 3674 IVs associated with 1502 plasma proteins.

Then subsequent to clumping at an LD R2 threshold of 0.4 to obtain IVs for each exposure (plasma protein) and calculated the IV strength for each protein to ensure that these IVs were a good genetic proxy for the corresponding plasma protein level (F-statistics were all greater than 20), we validate 3156 IVs associated with 1487 plasma proteins. Take IV-outcome dataset into consideration, we harmonize the IV-exposure data and IV-outcome data, we have 2965 IVs associated with 1422 plasma proteins. See Supplementary Material(IV selection and validation.xlsx) for more information.

### Proteome-wide MR analysis identified 15 plasma proteins significantly associated with MI risk

After primary MR analysis, 15 of these 1,422 proteins showed a significant genetically predicted causal association with MI risk (P −value < 3.510−5 (0.05/1,422))(Table 1). Considering that protein levels were rank-inverse normal transformed in the protein GWAS, for MR results, we paid more attention to the direction of the estimates than the magnitude of the estimates. Among these 15 significant proteins(Table 1), genetically predicted plasma levels of Plasminogen(Plasmin), Hepatocyte growth factor-like protein(MSP), Apolipoprotein B(Apo B), Transgelin-2(TAGLN2), Low-density lipoprotein receptor-related protein 4(LRP4), Complement C1s subcomponent(C1s), Angiostatin, Apolipoprotein C-III(Apo C-III), Proprotein convertase subtilisin/kexin type 9(PCSK9), Angiopoietin-related protein 4(ANGL4) were associated with higher MI risk, while the genetically predicted plasma levels of Fibronectin Fragment 4(FN1.4), Apolipoprotein B(Apo B), Interleukin-6 receptor subunit alpha(IL-6 sRa), Switch-associated protein 70(SWAP70), Fibronectin, Fibronectin Fragment 3(FN1.3) were associated with lower MI risk.

**Table 1.**
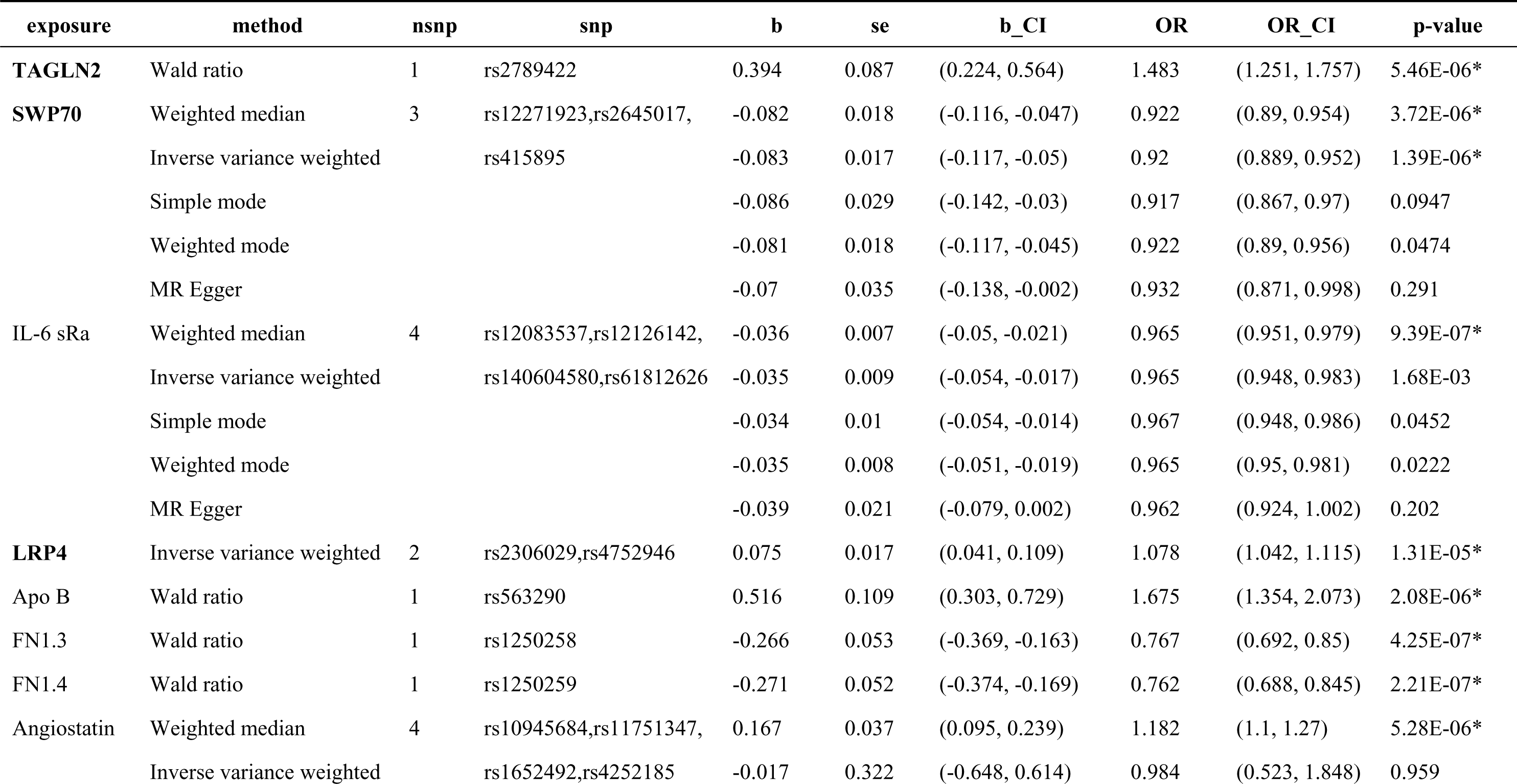
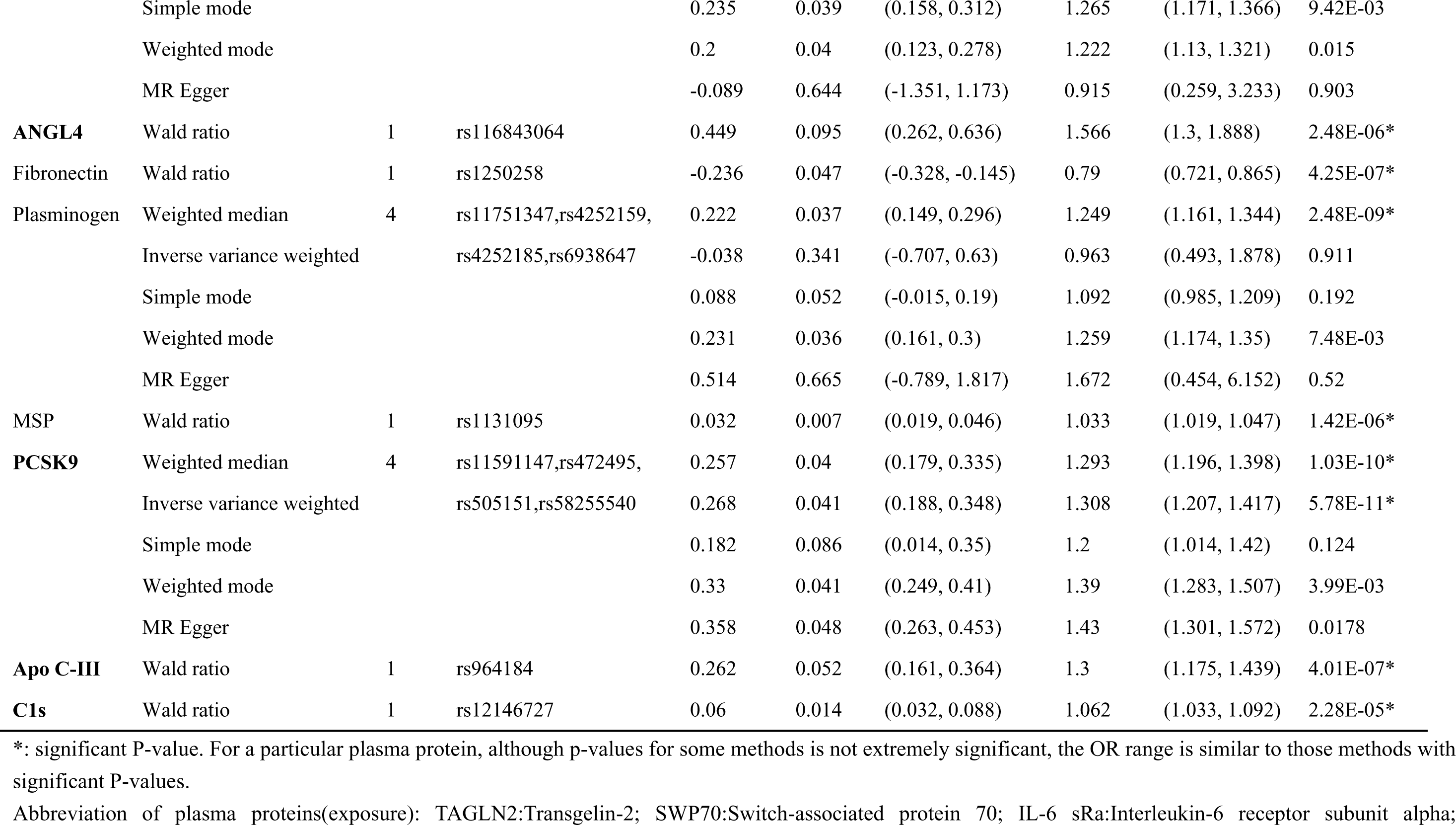
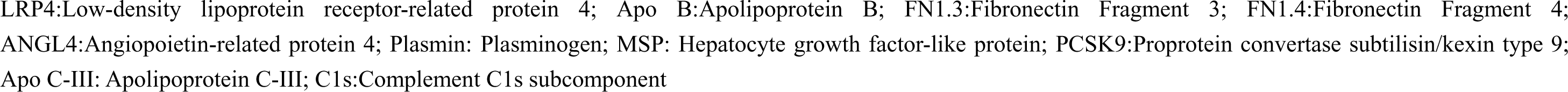
15 Plasma Proteins Identified Causally Associated with MI Risk.

| exposure | method | nsnp | snp | b | se | b_CI | OR | OR_CI | p-value |
| --- | --- | --- | --- | --- | --- | --- | --- | --- | --- |
| **TAGLN2** | Wald ratio | 1 | rs2789422 | 0.394 | 0.087 | (0.224, 0.564) | 1.483 | (1.251, 1.757) | 5.46E-06* |
| **SWP70** | Weighted median | 3 | rs12271923,rs2645017, | -0.082 | 0.018 | (-0.116, -0.047) | 0.922 | (0.89, 0.954) | 3.72E-06* |
|  | Inverse variance weighted |  | rs415895 | -0.083 | 0.017 | (-0.117, -0.05) | 0.92 | (0.889, 0.952) | 1.39E-06* |
|  | Simple mode |  |  | -0.086 | 0.029 | (-0.142, -0.03) | 0.917 | (0.867, 0.97) | 0.0947 |
|  | Weighted mode |  |  | -0.081 | 0.018 | (-0.117, -0.045) | 0.922 | (0.89, 0.956) | 0.0474 |
|  | MR Egger |  |  | -0.07 | 0.035 | (-0.138, -0.002) | 0.932 | (0.871, 0.998) | 0.291 |
| IL-6 sRa | Weighted median | 4 | rs12083537,rs12126142, | -0.036 | 0.007 | (-0.05, -0.021) | 0.965 | (0.951, 0.979) | 9.39E-07* |
|  | Inverse variance weighted |  | rs140604580,rs61812626 | -0.035 | 0.009 | (-0.054, -0.017) | 0.965 | (0.948, 0.983) | 1.68E-03 |
|  | Simple mode |  |  | -0.034 | 0.01 | (-0.054, -0.014) | 0.967 | (0.948, 0.986) | 0.0452 |
|  | Weighted mode |  |  | -0.035 | 0.008 | (-0.051, -0.019) | 0.965 | (0.95, 0.981) | 0.0222 |
|  | MR Egger |  |  | -0.039 | 0.021 | (-0.079, 0.002) | 0.962 | (0.924, 1.002) | 0.202 |
| **LRP4** | Inverse variance weighted | 2 | rs2306029,rs4752946 | 0.075 | 0.017 | (0.041, 0.109) | 1.078 | (1.042, 1.115) | 1.31E-05* |
| Apo B | Wald ratio | 1 | rs563290 | 0.516 | 0.109 | (0.303, 0.729) | 1.675 | (1.354, 2.073) | 2.08E-06* |
| FN1.3 | Wald ratio | 1 | rs1250258 | -0.266 | 0.053 | (-0.369, -0.163) | 0.767 | (0.692, 0.85) | 4.25E-07* |
| FN1.4 | Wald ratio | 1 | rs1250259 | -0.271 | 0.052 | (-0.374, -0.169) | 0.762 | (0.688, 0.845) | 2.21E-07* |
| Angiostatin | Weighted median | 4 | rs10945684,rs11751347, | 0.167 | 0.037 | (0.095, 0.239) | 1.182 | (1.1, 1.27) | 5.28E-06* |
|  | Inverse variance weighted |  | rs1652492,rs4252185 | -0.017 | 0.322 | (-0.648, 0.614) | 0.984 | (0.523, 1.848) | 0.959 |
|  | Simple mode |  |  | 0.235 | 0.039 | (0.158, 0.312) | 1.265 | (1.171, 1.366) | 9.42E-03 |
|  | Weighted mode |  |  | 0.2 | 0.04 | (0.123, 0.278) | 1.222 | (1.13, 1.321) | 0.015 |
|  | MR Egger |  |  | -0.089 | 0.644 | (-1.351, 1.173) | 0.915 | (0.259, 3.233) | 0.903 |
| **ANGL4** | Wald ratio | 1 | rs116843064 | 0.449 | 0.095 | (0.262, 0.636) | 1.566 | (1.3, 1.888) | 2.48E-06* |
| Fibronectin | Wald ratio | 1 | rs1250258 | -0.236 | 0.047 | (-0.328, -0.145) | 0.79 | (0.721, 0.865) | 4.25E-07* |
| Plasminogen | Weighted median | 4 | rs11751347,rs4252159, | 0.222 | 0.037 | (0.149, 0.296) | 1.249 | (1.161, 1.344) | 2.48E-09* |
|  | Inverse variance weighted |  | rs4252185,rs6938647 | -0.038 | 0.341 | (-0.707, 0.63) | 0.963 | (0.493, 1.878) | 0.911 |
|  | Simple mode |  |  | 0.088 | 0.052 | (-0.015, 0.19) | 1.092 | (0.985, 1.209) | 0.192 |
|  | Weighted mode |  |  | 0.231 | 0.036 | (0.161, 0.3) | 1.259 | (1.174, 1.35) | 7.48E-03 |
|  | MR Egger |  |  | 0.514 | 0.665 | (-0.789, 1.817) | 1.672 | (0.454, 6.152) | 0.52 |
| **MSP** | Wald ratio | 1 | rs1131095 | 0.032 | 0.007 | (0.019, 0.046) | 1.033 | (1.019, 1.047) | 1.42E-06* |
| **PCSK9** | Weighted median | 4 | rs11591147,rs472495, | 0.257 | 0.04 | (0.179, 0.335) | 1.293 | (1.196, 1.398) | 1.03E-10* |
|  | Inverse variance weighted |  | rs505151,rs58255540 | 0.268 | 0.041 | (0.188, 0.348) | 1.308 | (1.207, 1.417) | 5.78E-11* |
|  | Simple mode |  |  | 0.182 | 0.086 | (0.014, 0.35) | 1.2 | (1.014, 1.42) | 0.124 |
|  | Weighted mode |  |  | 0.33 | 0.041 | (0.249, 0.41) | 1.39 | (1.283, 1.507) | 3.99E-03 |
|  | MR Egger |  |  | 0.358 | 0.048 | (0.263, 0.453) | 1.43 | (1.301, 1.572) | 0.0178 |
| **Apo C-III** | Wald ratio | 1 | rs964184 | 0.262 | 0.052 | (0.161, 0.364) | 1.3 | (1.175, 1.439) | 4.01E-07* |
| **C1s** | Wald ratio | 1 | rs12146727 | 0.06 | 0.014 | (0.032, 0.088) | 1.062 | (1.033, 1.092) | 2.28E-05* |

\*: significant P-value. For a particular plasma protein, although p-values for some methods is not extremely significant, the OR range is similar to those methods with significant P-values.
Abbreviation of plasma proteins(exposure): TAGLN2:Transgelin-2; SWP70:Switch-associated protein 70; IL-6 sRa:Interleukin-6 receptor subunit alpha;
LRP4:Low-density lipoprotein receptor-related protein 4; Apo B:Apolipoprotein B; FN1.3:Fibronectin Fragment 3; FN1.4:Fibronectin Fragment 4; ANGL4:Angiopoietin-related protein 4; Plasmin: Plasminogen; MSP: Hepatocyte growth factor-like protein; PCSK9:Proprotein convertase subtilisin/kexin type 9; Apo C-III: Apolipoprotein C-III; C1s:Complement C1s subcomponent

### Sensitivity Analysis demonstrated the robustness of 7 candidate potential drug targets

#### Horizontal pleiotropy test

First, we performed a horizontal pleiotropy test to judge whether several SNPs associated with one protein has horizontal pleiotropy. According to Table 2, all the selected proteins passed the horizontal pleiotropy test with egger intercept apart from zero, indicating there exists no horizontal pleiotropy in the selected SNPs, fitting the assumption of Mendelian randomization.

**Table 2.**
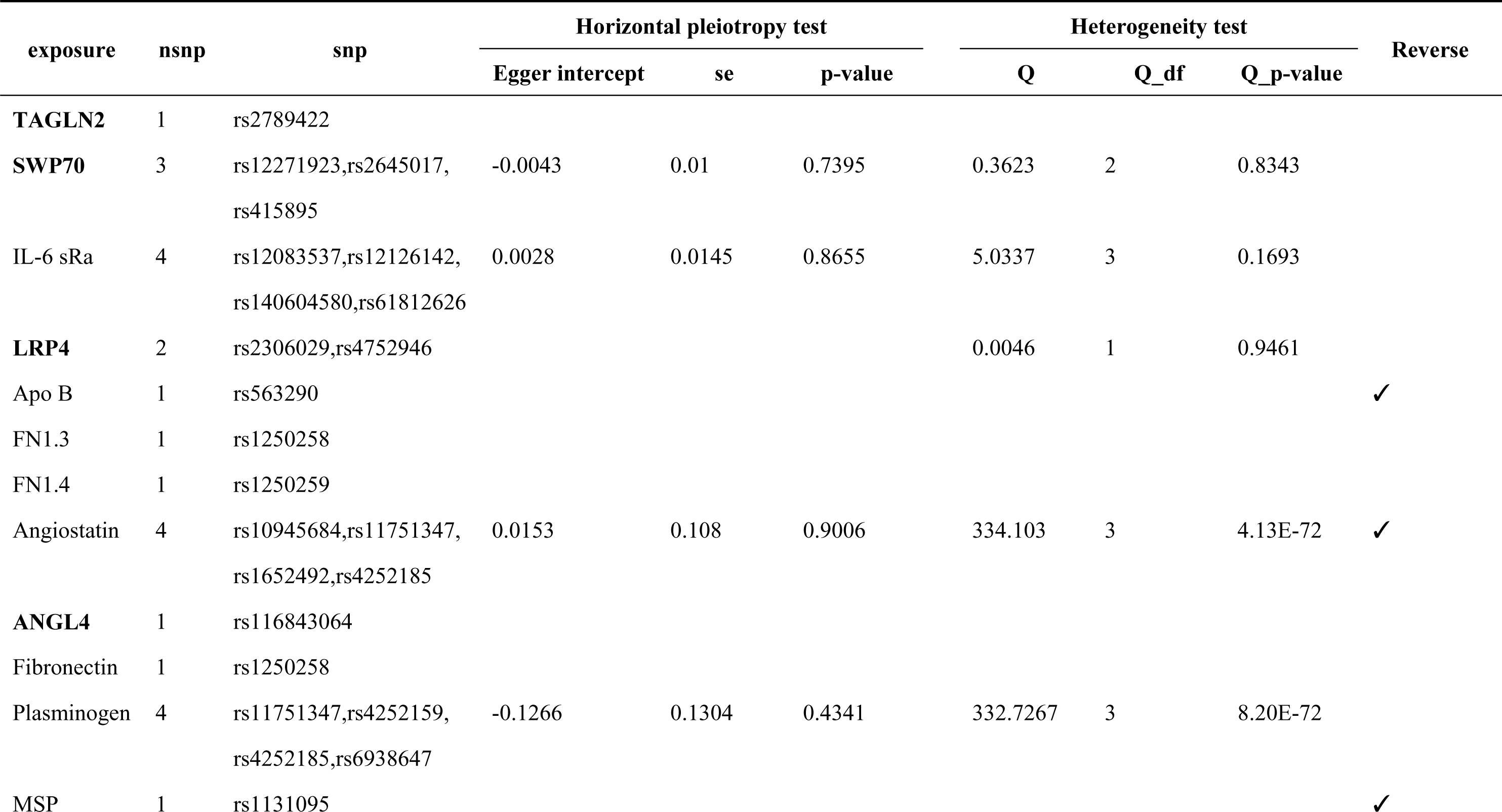
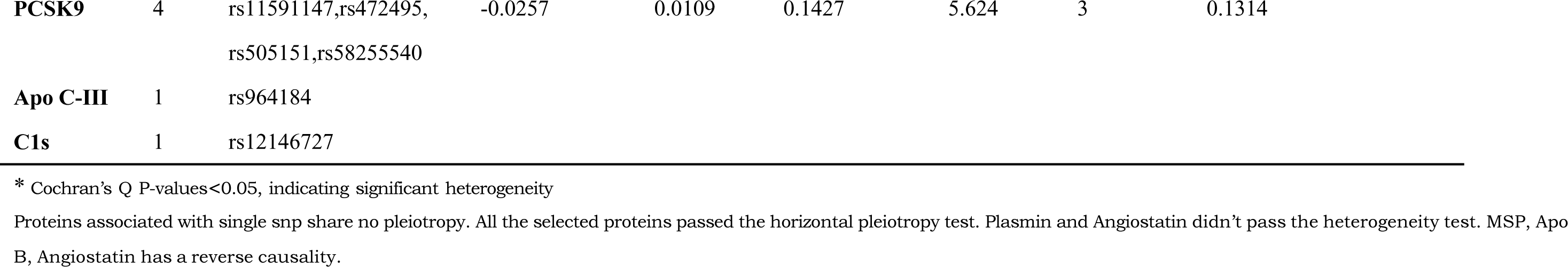
Sensitivity test of 15 candidate proteins.

| exposure | nsnp | snp | Horizontal pleiotropy test |  |  | Heterogeneity test |  |  | Reverse |
| --- | --- | --- | --- | --- | --- | --- | --- | --- | --- |
|  |  |  | Egger intercept | se | p-value | Q | Q_df | Q_p-value |  |
| **TAGLN2** | 1 | rs2789422 |  |  |  |  |  |  |  |
| **SWP70** | 3 | rs12271923,rs2645017,<br>rs415895 | -0.0043 | 0.01 | 0.7395 | 0.3623 | 2 | 0.8343 |  |
| IL-6 sRa | 4 | rs12083537,rs12126142,<br>rs140604580,rs61812626 | 0.0028 | 0.0145 | 0.8655 | 5.0337 | 3 | 0.1693 |  |
| **LRP4** | 2 | rs2306029,rs4752946 |  |  |  | 0.0046 | 1 | 0.9461 |  |
| Apo B | 1 | rs563290 |  |  |  |  |  |  | ✓ |
| FN1.3 | 1 | rs1250258 |  |  |  |  |  |  |  |
| FN1.4 | 1 | rs1250259 |  |  |  |  |  |  |  |
| Angiostatin | 4 | rs10945684,rs11751347,<br>rs1652492,rs4252185 | 0.0153 | 0.108 | 0.9006 | 334.103 | 3 | 4.13E-72 | ✓ |
| **ANGL4** | 1 | rs116843064 |  |  |  |  |  |  |  |
| Fibronectin | 1 | rs1250258 |  |  |  |  |  |  |  |
| Plasminogen | 4 | rs11751347,rs4252159,<br>rs4252185,rs6938647 | -0.1266 | 0.1304 | 0.4341 | 332.7267 | 3 | 8.20E-72 |  |
| MSP | 1 | rs1131095 |  |  |  |  |  |  | ✓ |

|  |  |  |  |  |  |  |  |  |
| --- | --- | --- | --- | --- | --- | --- | --- | --- |
| **PCSK9** | 4 | rs11591147,rs472495,<br>rs505151,rs58255540 | -0.0257 | 0.0109 | 0.1427 | 5.624 | 3 | 0.1314 |
| **Apo C-III** | 1 | rs964184 |  |  |  |  |  |  |
| **C1s** | 1 | rs12146727 |  |  |  |  |  |  |

\* Cochran's Q P-values<0.05, indicating significant heterogeneity
Proteins associated with single snp share no pleiotropy. All the selected proteins passed the horizontal pleiotropy test. Plasmin and Angiostatin didn't pass the heterogeneity test. MSP, Apo B, Angiostatin has a reverse causality.

#### Heterogeneity test

Secondly, we conducted a heterogeneity test sensitivity analysis. Our results revealed that proteins Plasmin and Angiostatin had Cochran’s Q P-values lower than 0.05. This suggests significant heterogeneity among these proteins, violating the assumption of Mendelian randomization. This warrants special consideration in subsequent analyses.

#### Reverse Causality Analysis

Thirdly, a reverse causality analysis was conducted, indicating that proteins MSP, Apo B, Angiostatin demonstrated reverse causality effects on MI. This could mean that myocardial infarction (MI) caused the expression of these plasma proteins, which, as originally thought. are not our founding potential targets for MI.

#### Colocalization Analysis

Lastly, in the colocalization analysis, at a threshold of 0.8, several proteins were found to share the same variants with MI(Table 3), including Apo B (rs12713844; rs563290), Transgelin-2(rs2789422), LRP4(rs2306029; rs4752946), C1s(rs12146727), Apo C-III (rs964184), PCSK9(rs11591147; rs472495; rs505151; rs58255540) and ANGL4(rs116843064). When the threshold was lowered to 0.7, additional variants in SWAP70 (rs10743116; rs12271923; rs2645017; rs415895) were also deemed to share the same variants with MI.

**Table 3.**
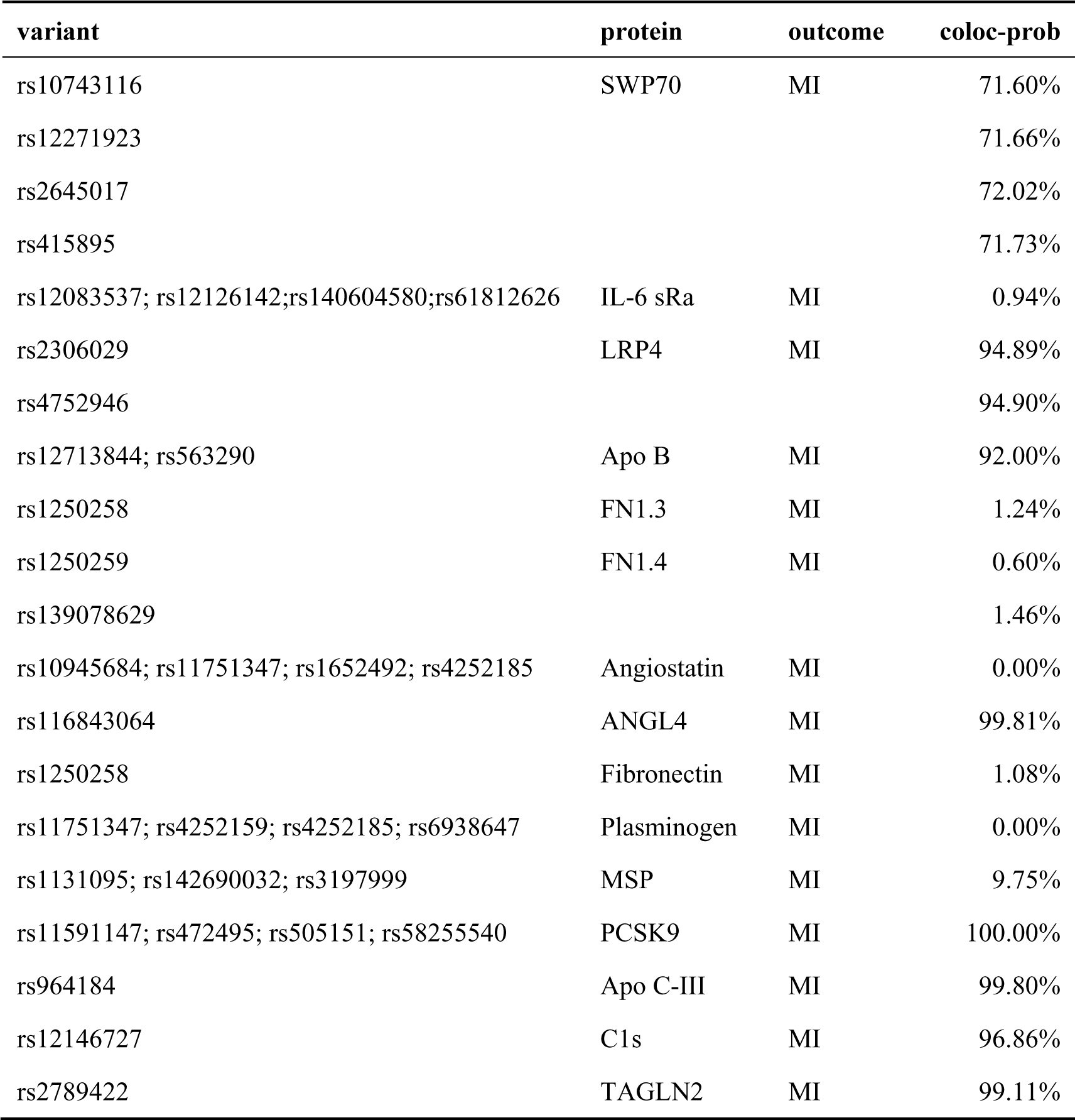
Colocalization Analysis.

Through this comprehensive set of analyses, we identified 7 candidate potential drug targets related to myocardial infarction (MI) more accurately, including SWP70, TAGLN2, LRP4, C1s, Apo C-III, PCSK9 and ANGL4, while also shedding light on possible false positives and false negatives. These findings will help deepen our understanding of the molecular mechanisms underlying MI and could have a positive impact on future drug development targeting this condition.

### Drugability assessment of potential drug targets

We searched DrugBank, PharmGKB, and TTD databases to fully evaluate the Drugability of our selected potential drug targets. According to the 3 databases, potential drug targets were classified into four categories: Successful Target (drugs targeting this protein have been successfully approved), Clinical trial Target (drugs targeting this protein are undergoing clinical trials), Literature-reported Target (this protein has been reported in the literature as a potential drug target), Not currently listed as druggable (no research using this protein as a drug target has been retrieved).(Table 4) In general, C1s is a successful target for Hereditary angioedema and Cold type autoimmune haemolytic anemia; Apo C-III, PCSK9 are Clinical trial Targets;TAGLN2 and ANGL4, are Literature-reported Target; LRP4 and SWP70 is Not currently listed as druggable.

**Table 4.** Drugability assessment of potential drug targets.

| Protein | Target Info | Target Name | Target type | Disease | Drugs |
| --- | --- | --- | --- | --- | --- |
| TAGLN2 | T93147 | Transgelin-2 (TAGLN2) | Literature-reported Target | not Available | not Available |
| SWP70 |  |  |  |  |  |
| LRP4 |  |  |  |  |  |
| Apo B | T07533 | Apolipoprotein B-100 (APOB) | Clinical trial Target | High blood cholesterol level | SPC4955 |
|  | T09507 | APOB messenger RNA (APOB mRNA) | Successful Target | Familial hypercholesterolemia; Hypercholesterolaemia | Mipomersen |
| ANGL4 | T98635 | Angiopoietin-related protein 4 (ANGPTL4) | Literature-reported Target | Not Available | Not Available |
| PCSK9 | T62206 | Proprotein convertase subtilisin/kexin type 9 (PCSK9) | Successful Target | Familial hypercholesterolemia; Chronic obstructive pulmonary disease;COPD | REGN-727 |
|  | T86652 | PCSK9 messenger RNA (PCSK9 mRNA) | Clinical trial Target | Cardiovascular disease; Metabolic disorder | MPSK3169A |
| Apo C-III | T86115 | ApoC-III messenger RNA (APOC3 mRNA) | Successful Target | Successful Target | Volanesorsen |
|  | T31518 | Apolipoprotein C-III (ApoCIII) | Clinical trial Target | Not Available | Not Available |
| C1s | T76024 | Complement C1s component (C1S) | Successful Target | Hereditary angioedema; Cold type autoimmune haemolytic anemia | Cinryze |

## Discussion

In the present study, we used proteome-wide GWAS data to conduct a Mendelian randomization analysis. After a series of stringent quality control measures to validate IVs, we obtained IVs that satisfied the three core assumptions of MR. Using these IVs, we identified 15 candidate plasma proteins (Plasminogen(Plasmin), Hepatocyte growth factor-like protein(MSP), Apolipoprotein B(Apo B), Transgelin-2(TAGLN2), Low-density lipoprotein receptor-related protein 4(LRP4), Complement C1s subcomponent(C1s), Angiostatin, Apolipoprotein C-III(Apo C-III), Proprotein convertase subtilisin/kexin type 9(PCSK9), Angiopoietin-related protein 4(ANGL4),Fibronectin Fragment 4(FN1.4), Apolipoprotein B(Apo B), Interleukin-6 receptor subunit alpha(IL-6 sRa), Switch-associated protein 70(SWAP70), Fibronectin, Fibronectin Fragment 3(FN1.3)) casually associated with MI. A full range of sensitivity analyses confirmed the robustness in the primary analysis, selecting 7 candidate plasma proteins including PCSK9,LRP4,APO C-III,ANGL4,SWP70,TAGLN2 and C1s. Finally, by searching multiple drug databases, we have a full understanding of the current status of drug development targeting these proteins, and suggest the possibility of multiple uses for approved drugs and new MI therapeutic targets.

### Protein targets associated with decrease of LDL-C

Elevated cholesterol levels, especially high levels of low-density lipoprotein cholesterol (LDL-C), increase the risk of atherosclerosis (also known as coronary heart disease). In this condition, cholesterol and other lipids accumulate on the walls of the blood vessels, forming “plaques”. These plaques can lead to arterial narrowing and hardening, restricting blood flow. When the coronary arteries (the main vessels supplying blood to the heart) are severely narrowed or blocked, the rupture of an unstable plaque followed by subsequent blood clot formation can trigger a myocardial infarction (heart attack). High LDL-C levels increase the risk of the formation and rupture of such unstable plaques. [35]

#### PCSK9

Proprotein convertase subtilisin/kexin type 9 (PCSK9) is A successful target for Familial hypercholesterolemia and Chronic obstructive pulmonary disease. The Low-Density Lipoprotein Receptor (LDLR) modulates blood cholesterol levels by binding to and clearing circulating LDL. It acts on low-density lipoprotein receptors (LDLR) and targets them for degradation, thus reducing the liver’s ability to remove LDL-cholesterol from the bloodstream, playing a crucial role in cholesterol metabolism. Inhibitors of PCSK9 are used to treat high levels of LDL-cholesterol. [36]

Several research reveals that the use of Proprotein Convertase Subtilisin/Kexin type 9 (PCSK9) inhibitors (monoclonal antibodies) can reduce LDL-C levels. Fourier et al.’s experiment [37] revealed that treatment with Evolocumab significantly reduced the risk of the composite outcome of cardiovascular death, MI, stroke, unstable angina, or coronary revascularization by 19%. The ODYSSEY outcome trial [38, 39] also showed similar results. Alirocumab significantly reduced the risk of the primary endpoint by 15% (HR 0.85, 95%CI 0.78–0.93, P = 0.003). In a recent meta-analysis of 67 randomized controlled trials (RCTs) [40] including 259,429 participants, PCSK9 inhibitors plus statin significantly reduced the risk of non-fatal MI (RR 0.82,95% CI 0.72– 0.93, p=0.003) or stroke (RR 0.74,95% CI 0.65–0.85, p<<0.001). This provides strong support for future drug research aiming at improving myocardial infarction outcomes by targeting this mechanism.

#### LRP4

LRP4 is not currently listed as druggable in the drug database, however its super family LRP5 and LRP6 are clinical trial Targets for solid tumour/cancer through Wnt/β-catenin signaling pathway [41, 42],where LRP4 also holds a pivotal position. Wnt/β-catenin signaling, a complex and highly conserved cell signaling system, plays a crucial role in the patterning, morphogenesis and growth of a variety of tissues and organs during development and in homeostasis in the adult, While Aberrant Wnt signaling activity is causally linked to degenerative diseases and cancers [43, 44], MI included. Three main phases, inflammation, angiogenesis and fibrosis [45, 46], widely recognized following MI, may exacerbate Wnt/β-catenin signaling and interfere with the balanced cardiac remodeling, thus these events could aggravate tissue damage and cause heart failure.

Recent research has found new therapeutic strategy to treat MI, using inhibitors of the canonical Wnt/β-catenin signaling pathway [47, 48],like anti-cancer drugs in patients after MI to prevent heart failure. Early using of these drugs in the first days after myocardial infarction can could inhibit Wnt signaling pathway, limit the loss of cardiomyocytes and fibrosis, thus promote myocardial regeneration, prevent adverse cardiac remodeling and improve LV function. [49] Dkk1 is one representative of the inhibitors which can enhance ischemic injury by promoting the internalization and elimination of basal LRP4/5.[50]

### Protein targets associated with balance of TG

#### Apo C-III

Apolipoprotein C-III (ApoCIII) is a clinical trial target, while its messenger RNA (APOC3 mRNA) has become a successful target for Hypertriglyceridemia and Hyperlipoproteinemia. This protein has its unique proatherogenic effect on metabolism dysfunction of TGs and other various lipid molecules, as well as its influence on acceleration of pro-inflammatory effects between monocytes and endothelial cells, causing the high risk of MI. [51] In a follow-up randomized controlled trial, Mendivil et al. [52] highlighted that the coronary heart disease risk posed by LDL is primarily due to the presence of apoC-III within the LDL.

In a retrospective case–control study, Gerber et al[53] investigated 90 consecutive patients with CHD and compared them with 209 hospital-based controls, finding an significant odds ratio in a cardiovascular event(OR=1.82,95% CI 1.18–2.79, p=0.003),in a non-HDL apoC-III (OR=2.44,95% CI 1.58–3.78, p¡0.001), HDL apoC-III(OR=0.64,95% CI 0.42–0.96, p=0.09), with each SD increase in total serum apoC-III.

#### ANGL4

ANGL4 is a Literature-reported Target. Lipoprotein lipase (LPL) hydrolyzes fatty acids from triglyceride-rich lipoproteins and regulates their distribution to peripheral tissues. ANGL4 mediates the inactivation of the lipoprotein lipase LPL [54, 55], thus playing a significant role in regulating serum triglyceride clearance and lipid metabolism. This has been confirmed by several human genome-wide association studies [56–59]. Numerous genetic studies indicate that elevated plasma triglycerides are associated with increased risk factors for cardiovascular disease [54, 55, 60]. Given ANGL4’s role in lipid regulation, ANGL4 is widely considered a potential drug target for cardiovascular diseases. It has been reported that plasma ANGL4 levels can predict future cardiovascular events, suggesting that ANGL4 might serve as a biomarker for coronary artery disease, including myocardial infarction [61], and an effective drug target.

### Protein targets associated with decrease of LDL-C

#### SWP70

SWAP 70 is not currently listed as a therapeutic target. Preliminary evidence from animal studies have shown that SWAP 70 may inhibit the development of non-alcoholic fatty liver disease (NAFLD) by suppressing lipid accumulation, inflammatory responses, and fibrosis. An ELISA study [35, 36] suggests that the SWAP-70 antibody may serve as a biomarker for the recurrence and prognosis of multiple sclerosis (MS). In the context of cardiovascular diseases, a gene knockout experiment [37] has shown that SWAP70 could inhibit the progression of cardiac hypertrophy, possibly through TAK1-dependent suppression of the mitogen-activated protein kinase signaling pathway. Lysosomes may also be involved in regulating the expression levels of SWAP70. Though its potential role in the prevention and treatment of myocardial infarction requires further investigation, these findings indicate a promising avenue for future research, especially in the realm of therapeutic applications.

### Protein targets with other mechanism

#### TAGLN2

TAGLN2 is a literature reported target. TAGLN2 is an actin-binding protein expressed in smooth muscle cells, related to the formation of the cytoskeletal structure, and plays a role in cell migration and tissue repair. Since smooth muscle cells and vascular repair have certain roles in the development of cardiovascular diseases, including myocardial infarction, it is theoretically possible that TAGLN2 may have an indirect association with cardiovascular diseases. Most studies on TAGLN2 focus on its relationship with non-cardiovascular diseases such as cancer, inflammation, and autoimmune diseases. Further research is needed to determine whether it can serve as a reliable biomarker and potential drug target for myocardial infarction.

#### C1s

C1s is a successful target. Complement system and C1s plays a critical role in maintaining homeostasis, which is associated with dysfunction and many disorders. The C1 complex, composed of C1q, C1r, and C1s (C1qr2s2), is the initiator of the classical complement activation pathway. At molecular level, C1r activates C1s so that it can, in turn, activate C2 and C4. C1s B chain is a serine protease that combines with C1q and C1r to form C1, which the first component of the classical pathway of the complement system. As C1s is activated under various pathological conditions and associated with the development of inflammation, autoimmunity and cancer, it has become an outstanding biomarker or target for diagnosis and treatment to large quantities of disease.

C1s has been a successful target for Hereditary angioedema and Cold type autoimmune haemolytic anemia, but remains literature-reported for MI. Complement system and C1s plays a critical role in maintaining homeostasis, which is associated with dysfunction and many disorders, including cardiovascular artery diseases like MI. [62] C1s is a member of the SCUBE1 family [63], which is a platelet granule protein that is exposed on the surface of platelets upon activation. Activated platelets are key factors in the development of arteriosclerosis. [64] Lin’s research [65] suggests that SCUBE may be a crucial platelet-endothelial adhesion molecule in the pathophysiology of cardiovascular disease development. As C1s is activated under various pathological conditions and associated with the development of inflammation, autoimmunity and cancer, it has become an outstanding biomarker or target for diagnosis and treatment to large quantities of disease. More researches can be conducted to use C1s as a target for MI.

### Our study has a lot of advantages

1. Comprehensive Data Analysis: By using an expansive dataset from the Icelandic Cancer Project and deCODE genetics, we were able to capture associations between a broad array of genetic variants and plasma proteins.
2. Robustness through MR Analysis: Mendelian Randomization allows for the identification of causal relationships, minimizing the impact of confounding factors typically encountered in observational studies.
3. Rigorous Sensitivity Analyses: Through heterogeneity tests, reverse causality analyses, and colocalization analyses, we added layers of validation to our findings, making our conclusions more robust.
4. Practical Implications: Our study does not merely identify genetic associations; it has tangible implications for the identification of potential drug targets, speeding up the typically long drug development process.

### However, our study still has some limitations

1. Potential Population Bias: Our study heavily relies on data from Icelandic individuals. While it provides a comprehensive dataset, the findings might not be generalizable to populations with different genetic backgrounds.
2. Incomplete Drugability Assessment: Though we explored drugability using prominent databases, emerging research or less-established potential drug targets might have been overlooked.
3. Complexity of MI Pathogenesis: MI is a multifactorial condition, and while our study sheds light on certain causal factors, there might be other significant variables or interactions not captured in our analysis.
4. Limitations of MR: Despite the strengths of MR, it’s based on several key assumptions, such as the relevance and exclusion criteria. If any of these are violated, it might introduce biases.

## Conclusion

Our study represents a significant stride in understanding the causal relationships between plasma proteins and MI risk. By harnessing the power of Mendelian Randomization and extensive genetic datasets, we’ve pinpointed promising drug targets for MI, offering a beacon of hope for streamlining future therapeutic interventions. However, as with all research, our findings have certain limitations. Further research in diverse populations and incorporating more extensive drugability datasets will be essential to validate and expand upon our conclusions.

## Supplementary information

IV selection and validation.xlsx is result of IV selection and validation.

## Data Availability

https://www.decode.com/summarydata

https://www.nature.com/articles/s41588-021-00978-w#MOESM4

https://academic.oup.com/eurheartj/article/42/9/919/6126843#supplementary-data

http://db.idrblab.net/ttd/

https://www.pharmgkb.org/

https://go.drugbank.com/

https://www.ebi.ac.uk/gwas/studies/GCST011365

## Non-standard Abbreviations and Acronyms

MI: myocardial infarction
FDA: Food and Drug Administration
GWAS: genome-wide association study
IV: instrumental variable
IVW: inverse variance weighted method
LD: linkage disequilibrium
MAF: minor allele frequency
MHC: major histocompatibility complex
MR: mendelian randomization
pQTL: quantitative trait locus
SNP: single nucleotide polymorphism
TTD: therapeutic target database

